# Evaluation of the PREDIGT Score in Discriminating Parkinson Disease from Neurological Health

**DOI:** 10.1101/2020.11.29.20240028

**Authors:** Juan Li, Tiago A. Mestre, Brit Mollenhauer, Mark Frasier, Julianna J. Tomlinson, Claudia Trenkwalder, Tim Ramsay, Douglas Manuel, Michael G. Schlossmacher

**Affiliations:** Neuroscience Program, Ottawa Hospital Research Institute; Clinical Epidemiology Program, Ottawa Hospital Research Institute; Methods Centre, Ottawa Hospital Research Institute; Division of Neurology, The Ottawa Hospital, Ottawa, ON., Canada; Division of General Medicine, Department of Medicine, The Ottawa Hospital, Ottawa, ON., Canada; University of Ottawa Brain and Mind Research Institute; Department of Medicine, Faculty of Medicine, University of Ottawa; Ottawa, ON., Canada; Department of Cellular and Molecular Medicine, Faculty of Medicine, University of Ottawa; Ottawa, ON., Canada; Elena-Paracelsus Klinik, University of Goettingen, Kassel, Germany; Michael J. Fox Foundation for Parkinson’s Research, New York, USA

## Abstract

**Background:** We previously created the PREDIGT Score as an algorithm to predict the incidence of Parkinson disease. The model rests on a hypothesis-driven formula, [P_R_=(E+D+I)xGxT], that uses numerical values for five categories known to modulate Parkinson’s risk (P_R_): environmental exposure (E); DNA variants (D); evidence of gene-environment interactions (I); gender (G); and time (T). Notably, the formula does not rely on motor examination results.

**Methods:** To evaluate the PREDIGT Score, we tested it in two established case-control cohorts: ‘De Novo Parkinson Study’ (DeNoPa) and ‘Parkinson’s Progression Marker Initiative’ (PPMI). Using baseline data from 589 patients and 309 controls enrolled in the DeNoPa and PPMI cohorts, we evaluated the PREDIGT Score’s discriminative performance in distinguishing Parkinson’s patients from healthy controls by area-under-the-curve (AUC) analyses.

**Findings:** When examining cohorts separately and using all available variables in each cohort to calculate the PREDIGT Score, AUCs were 0.83 (95% CI 0.77-0.89) for DeNoPa and 0.87 (95% CI 0.84-0.9) for PPMI, respectively, in distinguishing Parkinson disease patients from healthy individuals. When combining DeNoPa and PPMI data sets by using eleven variables that had been collected in both cohorts, the PREDIGT Score discriminated patients from controls with an AUC of 0.84 (95% CI 0.81-0.87). The mean score of Parkinson disease patients was significantly higher than that of control individuals at 108.48 (±52.08) and 47.33 (±34.1), respectively (p < 0.0001).

**Interpretation:** Our results demonstrate a robust performance of the original PREDIGT Score in distinguishing patients diagnosed with Parkinson disease from neurologically healthy subjects without reliance on motor examination data. In future efforts, the predictive performance of the algorithm will be studied in longitudinal cohorts of at-risk persons.

**Funding:** Parkinson Canada, Michael J. Fox Foundation, Department of Medicine (The Ottawa Hospital), Uttra & Subash Bhargava Family, Paracelsus-Elena-Klinik Kassel, Parkinson Fond Deutschland, and Deutsche Parkinson Vereinigung.

## INTRODUCTION

Parkinson disease (PD) remains an incurable neurodegenerative condition. The worldwide prevalence of PD is 6.1 million^1^ and its incidence is expected to double by 2030.^2^ As of 2020, no disease-modifying agents have been approved and no preventive therapy exists to delay the development of clinical PD. The ability to reliably identify a population at higher risk of PD could facilitate the design of trials aimed at an early intervention, such as by modifying known risk factors.

Predicting the onset of PD in neurologically healthy individuals is challenging for two reasons: one, the incomplete understanding of the disease’s aetiology; and two, the lack of sufficiently large, prospective cohorts of neurologically healthy participants followed for the development of PD. Recently, several models have been developed to predict onset and progression of PD using data analyses-driven and Bayesian classifier-based approaches. Berg *et al*. developed a three-step approach with pre-screening for age, followed by primary screening for positive family history and/or hyposmia, and secondary screening for the presence of augmented *S. nigra* echogenicity, as established by transcranial ultrasound. For every 16 positively screened subjects, one developed PD within three years of follow-up, which corresponded to a sensitivity of 80.0% (95% CI 44.5%-96.5%), a specificity of 90.6% (95% CI 90.4%-90.7%), and a positive predictive value of 6.1% (95% CI 3.4%-7.3%).^3^

A classifying model developed by Nalls *et al*. included the assessment of olfactory function, calculation of genetic risk by selecting 30 distinct DNA markers, a family history of PD, age, and sex. The composite score generated by stepwise logistic regression analysis distinguished PD patients from healthy controls (HC) in the ‘Parkinson’s Progression Marker Initiative’ (PPMI) cohort at an area-under-the-curve (AUC) value of 0.92 (95% CI 0.90-0.95).^4^ External validation of this model showed good classification of PD with AUCs ≥ 0.9.

The recently proposed model by Schrag *et al*., which was based on backward multivariate logistic regression analysis, showed discriminative power between PD and control subjects of an AUC of 0.80 (95% CI 0.78-0.81).^5^ In their approach, neurological and psychiatric variables, such as tremor, rigidity, difficulty with balance, cognitive decline and depression, were added to the presence of autonomic variables, which included constipation, hypotension, hypersalivation and urinary dysfunction.^5^

In contrast to these previous studies that relied on statistical methods to develop prediction rules for PD, the PREDIGT Score is a hypothesis-driven model.^6^ We previously postulated that the development of ‘idiopathic’ PD results from contributions by five risk categories: <u>E</u>xposome (E); genetic susceptibility (<u>D</u>NA variants; D); lasting tissue responses stemming from gene-environment Interactions (I); sex/<u>G</u>ender (G); and age, as in the passage of <u>T</u>ime (T). We had further hypothesized that from these risk elements a final score can be calculated by assigning values to each category, as computed by the formula: P_R_=(E+D+I)xGxT.^6^ Of note, for some risk categories, where details of variables could be incomplete, such as for an individual’s microbial exposure history within his/her nasal cavity (E) and/or regarding genetic polymorphisms (D), we had chosen surrogates, such as hyposmia and family history, respectively.^6^ Of importance, motor assessment-based data, as gleaned from routine neurological examinations, were not included in the original PREDIGT model.

The variables from the five categories (E, D, I, G, T) were selected and assigned with concrete values based on: (1) the hypothesis that PD is a complex disorder that requires more than one risk element in its aetiology; (2) an umbrella review of 75 meta-analyses to determine the effect size of individually selected variables; and (3) a model for pathogenesis that sees a gradual evolution from a healthy state to a prodromal phase on to the manifestation of cardinal signs, which result in the diagnosis of PD.^7^ A related objective was to develop and validate a score to predict future PD incidence rates in neurologically still healthy individuals.^6^

As an initial step, we set out to evaluate the discriminative ability of the PREDIGT Score to distinguish persons that have already been diagnosed with PD from neurologically healthy controls. To this end, we analyzed data from two well established, previously characterized and published case-control studies, *i*.*e*., the ‘De Novo Parkinson Study’ (DeNoPa)^8^ and PPMI.^9^

## METHODS

### Source of data and participants

We used de-identified baseline data of DeNoPa and PPMI. The DeNoPa cohort^8^ is a single-center, observational, longitudinal, Germany-based study of patients with a recent (“*de novo*”) diagnosis of PD (UK Brain Bank Criteria^10^) naïve to L-DOPA at baseline and of ageas well as sex-matched, neurologically healthy controls. Exclusion criteria included previously known or subsequently detected (during longitudinal follow up) conditions, such as normal-pressure hydrocephalus, cerebrovascular disease, signs or symptoms of atypical PD (*e*.*g*., multisystem atrophy or progressive supranuclear palsy), and medication-induced parkinsonism. Healthy controls (HC) were recruited through friends and relatives of enrolled PD subjects, other patients of the hospital-based clinic as well as through newspaper advertisement in early 2009. Controls had to be without any active, known or previously treated condition of their central nervous system, which included psychiatric diseases.

The original PPMI study (PPMI-1)^9^ is an international, multi-centric (n=33), observational, longitudinal study. PD patients were untreated and diagnosed within the previous two years. A positive dopamine transporter (DAT) scan, as proven by DAT SPECT imaging, was a study eligibility criterion. In addition, individuals with other movement disorders were excluded, as described above for DeNoPa participants. Healthy controls had to have no significant neurological dysfunction, no first-degree family member of PD, and a Montreal Cognitive Assessment (MoCA) test score of 26-30 (maximum score 30).

The study was approved by local ethics boards at all participating clinical sites within the DeNoPa and PPMI cohorts; analyses of de-identified data were approved by the Regulatory Ethics Board of The Ottawa Hospital (20180010-01H).

### Procedures and statistical analyses

**Figure 1** illustrates a summary of our workflow. The published original PREDIGT Score included 30 variables associated with altered risk for developing PD.^6^ Among those, 20 variables were captured between the two study cohorts (DeNoPa, n=19; PPMI, n=14). After excluding 4 variables with free-text data entries (*e*.*g*., infection and inflammation history) and low prevalence of exposure (*e*.*g*., MPTP-neurotoxin exposure, known encephalitis), we included 16 variables (as listed in **Figures 2, 4**, and **Suppl. Table 1**). Most variables had a low rate of missing data (<5%); we used predictive mean matching method for imputation of missing data.^11,12^ Cerebrospinal fluid (CSF) biomarker data for total α-synuclein protein and total tau protein concentrations and genetic risk scores (GRS) had higher missing data rates and could not be reliably imputed.

**Table 1:** Demographic characteristics of adults enrolled in the single-centre DeNoPa and multi-centric PPMI cohorts

| .. | DeNoPa |  |  | PPMI |  |  |
| --- | --- | --- | --- | --- | --- | --- |
|  | Case | Control | p | Case | Control | p |
| Characteristic | n (%) = 159 (59.1) | n (%) = 110 (40.9) | .. | n (%) = 430 (68.4) | n (%) = 199 (31.6) | .. |
| Male | 105 (66.04) | 67 (60.91) | 0.4 | 282 (65.58) | 128 (64.32) | 0.8 |
| Age at baseline (years) | 66 (40-85) | 65.5 (44-84) | 0.6 | 62 (34-85) | 62 (31-84) | 0.4 |
| Male | 66 (40-85) | 67 (44-84) | 0.8 | 63 (35-85) | 63 (31-83) | 0.7 |
| Female | 67.5 (41-84) | 64 (51-74) | 0.3 | 61 (34-82) | 60 (31-84) | 0.4 |
| 10-year age brackets | .. | .. | .. | .. | .. | .. |
| <40 | 0 (0) | 0 (0) | .. | 13 (3.02) | 9 (4.52) | .. |
| 40–50 | 8 (5.03) | 2 (1.82) | .. | 37 (8.6) | 20 (10.05) | .. |
| 50–60 | 38 (23.9) | 22 (20) | .. | 116 (26.98) | 54 (27.14) | .. |
| 60–70 | 52 (32.7) | 57 (51.82) | .. | 168 (39.07) | 73 (36.68) | .. |
| 70–80 | 54 (33.96) | 28 (25.45) | .. | 88 (20.47) | 37 (18.59) | .. |
| 80+ | 7 (4.4) | 1 (0.91) | .. | 8 (1.86) | 6 (3.02) | .. |

Data are n (%), or median (min–max). DeNoPa=De Novo Parkinson Study. PPMI=Parkinson’s Progression Marker Initiative.

**Figure 1:**
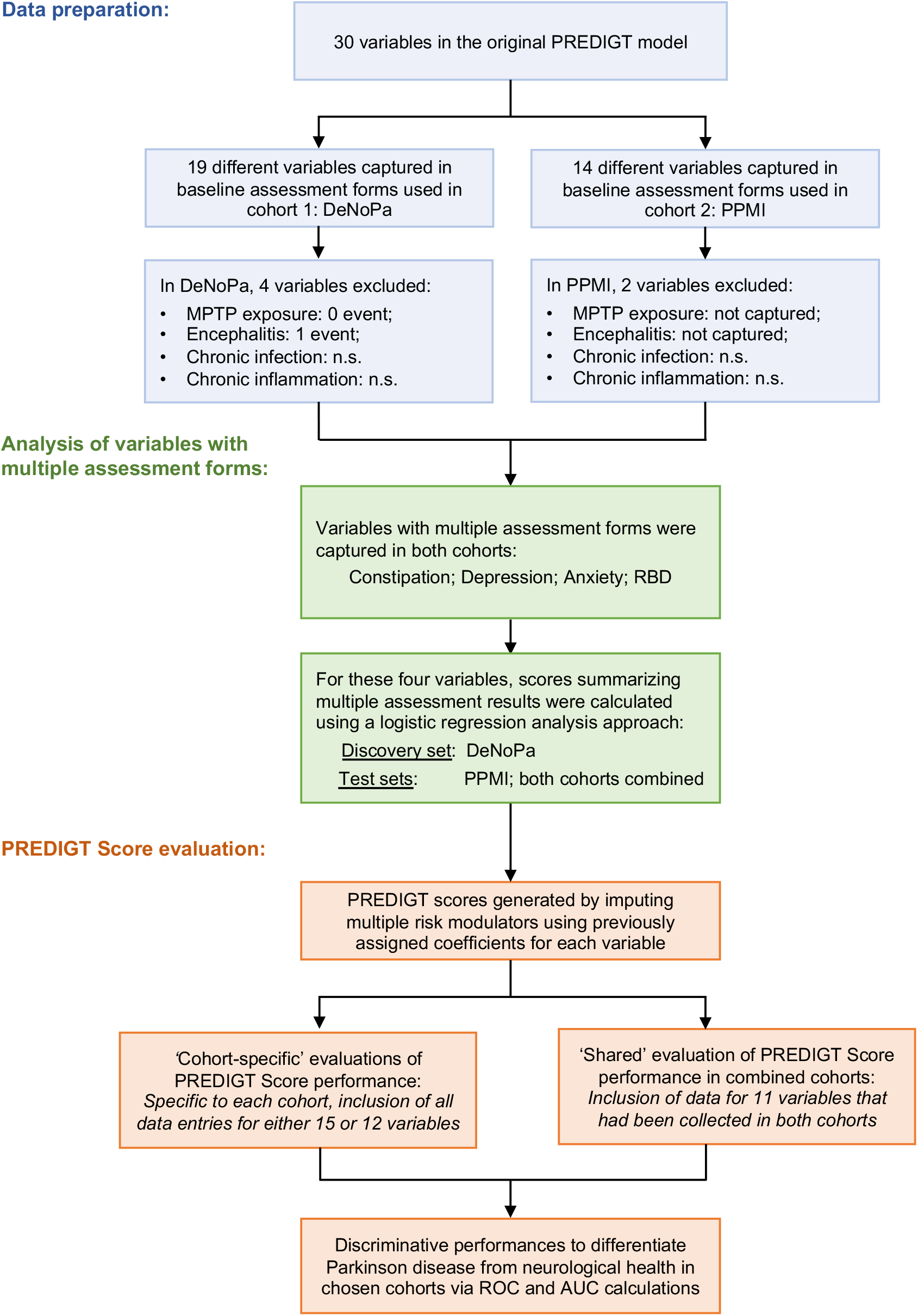
Workflow of this study. n.s.=not standardized. DeNoPa=De Novo Parkinson Study. PPMI=Parkinson’s Progression Marker Initiative. RBD=REM Sleep Behavior Disorder.

**Figure 2:**
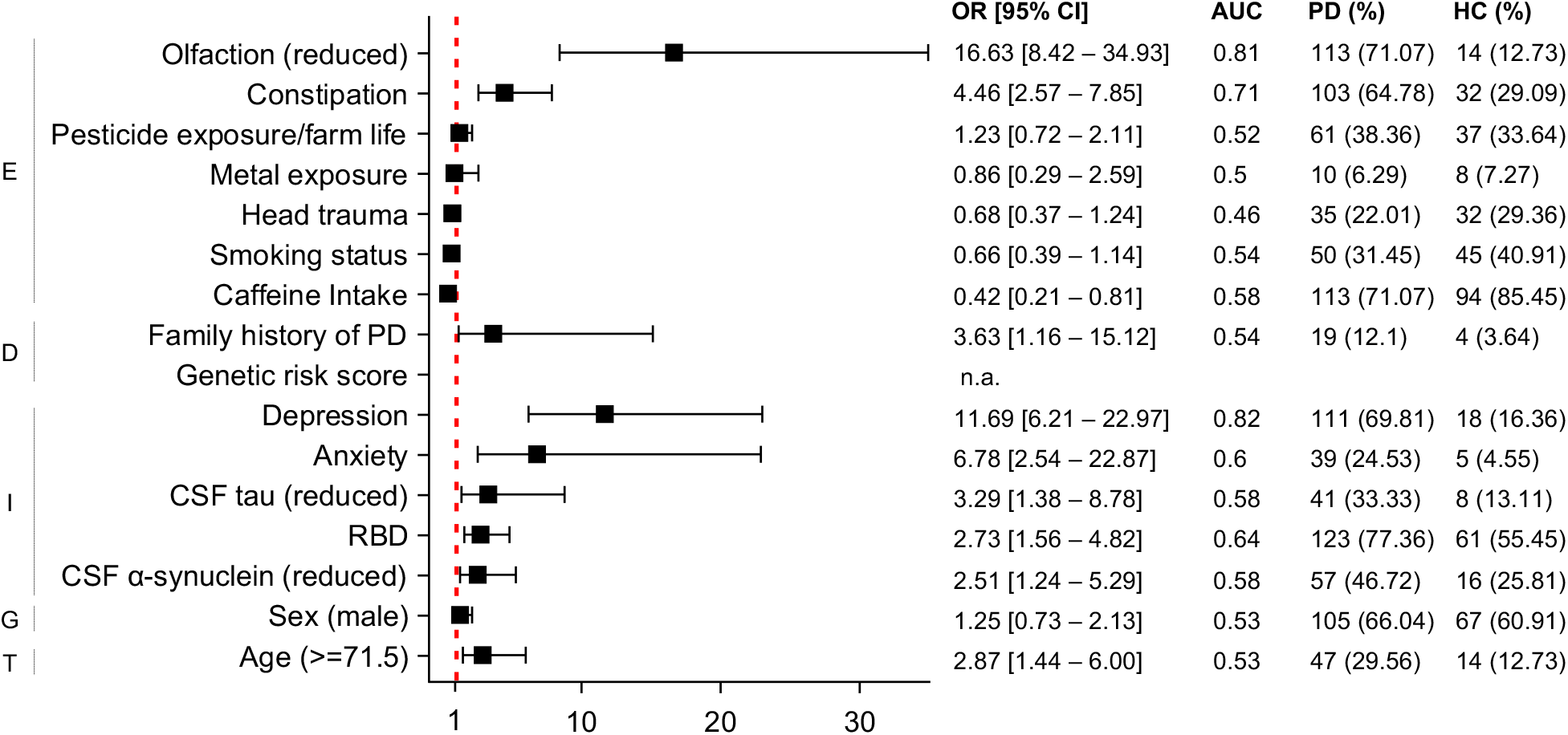
Effect sizes of fifteen selected variables in the DeNoPa cohort. Overall effect sizes for each variable, as summarized by the OR [95% CI], the AUC values from corresponding univariate logistic regression analyses, and the numbers and percentages (%) of positive cases in the PD and HC groups. The cut off value for age (*i*.*e*., 71.5 years of age) corresponds to the maximum Youden Index to best differentiate PD from HC subjects. n.a.=not available. DeNoPa=De Novo Parkinson. PD=Parkinson disease. HC=healthy controls. OR= odds ratio. CI=confidence interval. ROC=receiver operating characteristic. AUC=area under the ROC curve. RBD=REM Sleep Behavior Disorder. CSF=Cerebrospinal fluid.

Therefore, only subjects with complete data sets after their imputation were included: 33.3% of originally enrolled DeNoPa participants were excluded due to incomplete α-synuclein and/or tau data; 12.1% of originally enrolled PPMI participants were excluded due to incomplete α-synuclein, tau, and/or GRS data. Demographic information (such as sex, age) did not show significant differences between included subjects *vs*. all other individuals, except for healthy controls in the DeNoPa cohort, who had a higher proportion (47.4%) opting out of lumbar puncture tests to collect CSF compared with the PD group (23.5%).

### Assessments

Multiple instruments to evaluate non-motor symptoms were used in DeNoPa and PPMI (for details of screening instruments and their abbreviations, see **Suppl. Table 1**): in DeNoPa, recent non-motor symptoms (both severity and frequency) were self-reported using the MDS-UPDRS I, Scopa-AUT, PD NMSS, and PD NMS questionnaires. MDS-UPDRS I and Scopa-AUT were also used in PPMI for assessing the non-motor aspects of PD. Questionnaires for depression (GDS, BDI, MADRS), anxiety (STAI), and REM sleep behavior (RBD SQ) were also included in at least one of the two cohorts.^8, 9^ Sniffin’ Sticks-16 test and UPSIT were used in DeNoPa and PPMI, respectively, to quantify olfactory function. We classified patients as having normosmia, hyposmia, or anosmia by using published cut off values.^13,14^ In PPMI, participants’ inherited risk was summarized using a PD-specific genetic risk score (GRS), as generated from 30 markers implicated and replicated in one or more studies.^4^ Genetic data of DeNoPa participants are currently under analysis at the University of Lubeck; hence, family history of PD was used as a surrogate for missing genetic risk information. Different types of ELISA platforms were used to quantify the total tau concentration in these two cohorts. CSF biomarkers (for total α-synuclein and total tau) were therefore transformed for each subject into percent rank to ensure that data in these two cohorts were comparable.

### Data Analyses

Variables such as constipation, depression, anxiety, and REM sleep behaviour disorder (RBD) were assessed using multiple tools. Assessments that were captured in both cohorts were selected for further analysis (see **Suppl. Table 1**). There, multivariate logistic regression was applied to DeNoPa as the discovery set to generate summary scores of each variable separately (for details, see **Suppl. Table 2**). Optimal cut-off values, based on the maximum of Youden Indices,^15^ were identified to best distinguish PD patients from healthy controls. For external validation, the same models and cut-off values were applied to PPMI, and subsequently, to the combined cohort. The individual contribution of other, independent predictors was assessed using calculations for odds ratios and AUCs from univariable logistic regression analyses and illustrated by forest plots (**Figures 2, 4**). Based on the criteria shown in **Suppl. Table 1**, most variables were transformed into a binary format (positive/negative) or ordinal format with multiple levels.

The PREDIGT Score of each participant was calculated by adding previously assigned values^6^ given the status of corresponding variables, as summarized in **Suppl. Table 1**. Important for this study, these parameter values were not derived from the PPMI or DeNoPa data sets.

The discriminative performance of the PREDIGT Score was evaluated in three scenarios: 1) The formula was first tested using the DeNoPa cohort, which captured most of the variables that were selected in the original PREDIGT Score. 2) The PPMI cohort was then used for further evaluation. Although some PREDIGT variables were not captured in the current PPMI study protocol, sufficient results for each category of the original model were available. 3) The third evaluation was a ‘shared variable’-based analysis, in which participants of the two cohorts were combined. There, only variables captured in clinical research forms (CRFs) for both cohort studies were used (**Figure 1**; see also variables shaded in pink in **Suppl. Table 1**). Note, the pooling of DeNoPa and PPMI was made possible because participants in these cohorts had similar clinical and demographic characteristics (**Table 1**).

The model was assessed for overall discriminative performance using receiver operating characteristic (ROC) curves and calculations of corresponding AUC values. Reported sensitivity and specificity were relative to the optimal threshold identified using the Youden Index. Score density plots and box plots were used to illustrate the degree of distinction between the scores of PD *vs*. HC subjects.

Statistical analyses were performed using ‘R’ (version 3.6.0). Library ‘rms’ was used for logistic regression; ‘Hmisc’ was used for data imputation; forest plots were plotted using Prism 8.0, other plots were generated using ‘ggplot2’ in R.

## RESULTS

In the initial analysis, we included 269 DeNoPa subjects (PD: n=159; HC: n=110) and 629 PPMI individuals (PD: n=430; HC: n=199). Generally, DeNoPa and PPMI participants had comparable clinical and demographic characteristics (**Table 1**). PPMI had a higher percentage of PD patients (68.4%) than did DeNoPa (59.1%). The sex ratio was similar across cohorts and disease categorization: approximately two thirds of participants in each group were males. Median ages and their ranges were comparable across cohort, sex and disease categorization (PD, HC), although participants in DeNoPa were slightly older than those in PPMI (4 years difference in median age) and males were slightly older than females, all statistically insignificant (p>0.05). In both cohorts, we focused on variables previously identified as potentially informative **(Suppl. Table 1**).^6^ Data points related to motor performances were not included in the analysis of the PREDIGT Score.

### DeNoPa cohort

Variables that were included in this evaluation and their individual effect size of distinguishing PD from healthy controls are listed in **Figure 2**. Many associations between variables within PREDIGT categories and PD, such as hyposmia, constipation, caffeinated beverage intake, family history of PD, depression, anxiety, RBD, and reduction in CSF markers (total α-synuclein and total tau) were statistically significant, as expected from the literature.^6^ Among individual variables, the presence of depression (AUC: 0.82), hyposmia (AUC: 0.81), and constipation (AUC: 0.71) were particularly informative in distinguishing PD from HC (**Figure 2**). This univariate analysis supported the assumption stated for the original PREDIGT Score that these variables within factors ‘E’ and ‘I’ were associated with an altered overall risk of developing PD. Category ‘D’, where family history was a surrogate for genomic risk, was less discriminative in the DeNoPa cohort.

As illustrated in the score distribution plot (**Figure 3 (a)**) for the DeNoPa cohort, the HC group (n=59) had a lower PREDIGT Score compared with PD patients (n=120). The mean score (±SD) was significantly (p < 0.0001) lower than that of the PD group at 38.46 (40.83) and 101.84 (53.31), respectively (**Figure 3 (b)**). The AUC value for the PREDIGT Score in the DeNoPa cohort was 0.83 (95% CI 0.77-0.89) (**Figure 3 (c)**). Red dashed lines in **Figure 3 (a)** and **(b)** indicate the optimal threshold (49.2) that corresponds to the maximum value of the Youden Index. Using 49.2 as the threshold, the PREDIGT Score could distinguish PD patients from HC subjects in the DeNoPa cohort with 0.88 sensitivity (95% CI 0.81-0.93) and 0.68 specificity (95% CI 0.56-0.8).

**Figure 3:**
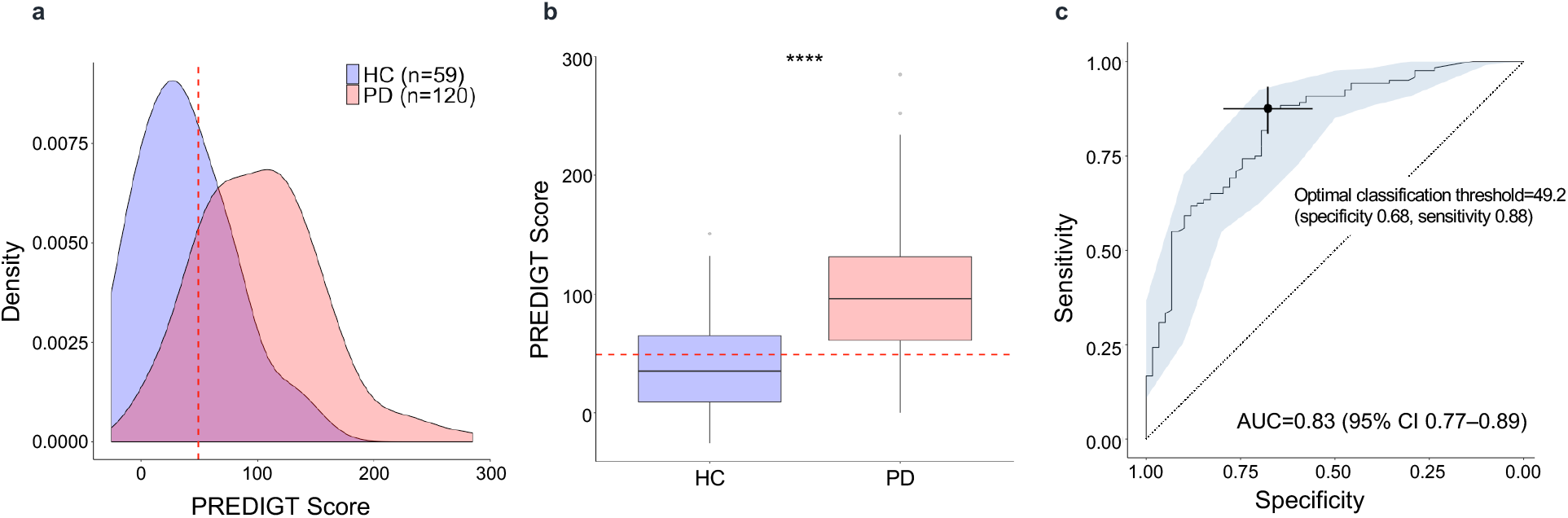
Cohort-specific evaluation of the PREDIGT Score performance using the DeNoPa cohort. (a) Score density plot; (b) box plot; and (c) ROC curve. The colour blue represents the HC group, pink represents PD patients. The mean score (with standard deviation) of healthy controls at 38.46 (±40.83) was significantly lower when compared to the PD group at 101.84 (±53.31) (p<0.0001). Red, dashed lines in panels (a), (b) and the cross in (c) indicate the optimal threshold (score, 49.2) to distinguish between PD and HC groups corresponding to the maximum Youden Index (specificity, 0.68; sensitivity, 0.88). Light blue shading in the ROC curve indicates the bootstrapestimated 95% CI within the AUC. DeNoPa=De Novo Parkinson. PD=Parkinson disease. HC=healthy controls. ROC=receiver operating characteristic. AUC=area under the ROC curve. CI=confidence interval.

**Figure 4:**
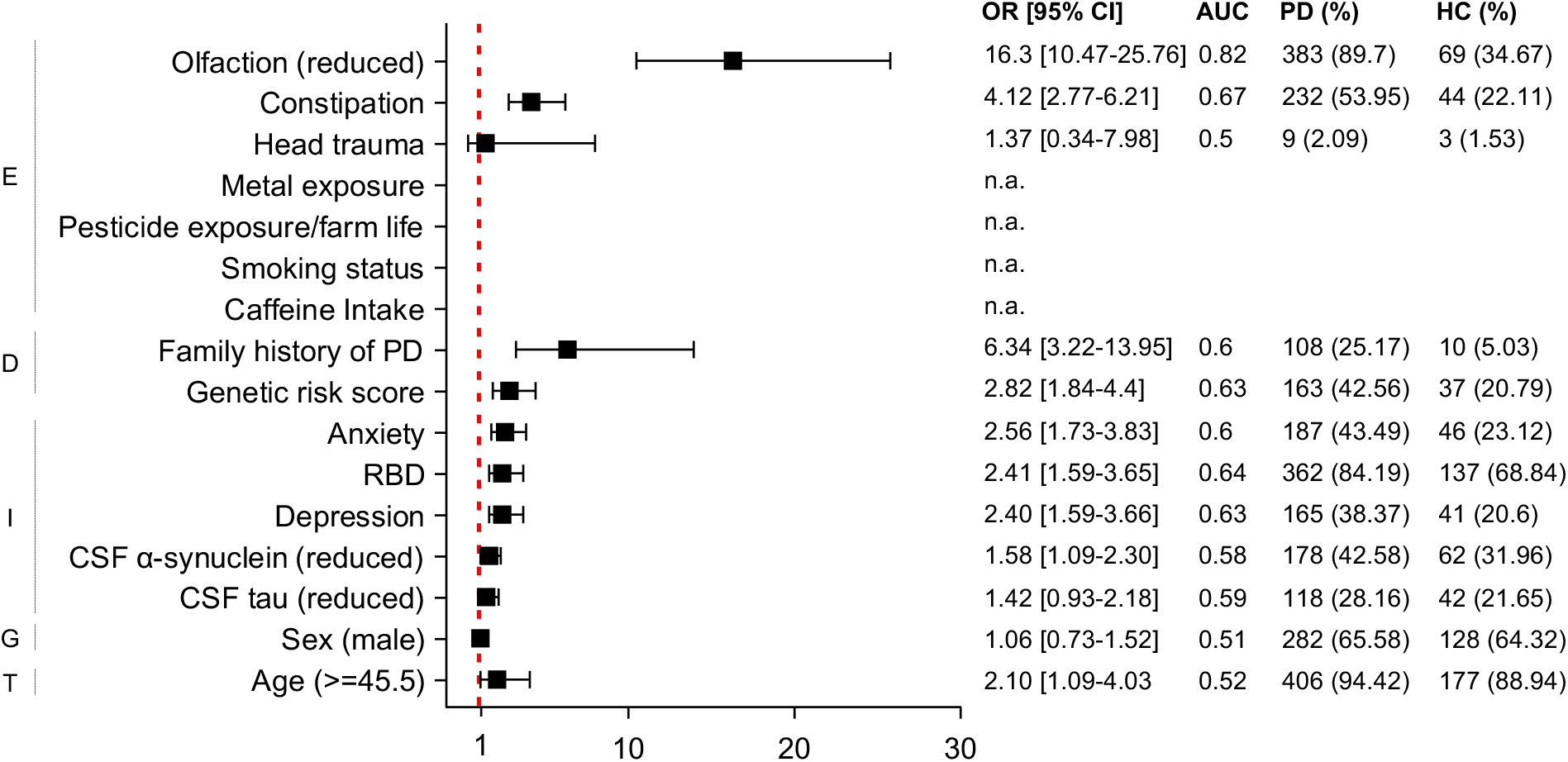
Effect sizes of twelve selected variables in the PPMI cohort. Overall effect sizes for each variable, as summarized by the OR [95% CI], the AUC values from corresponding univariate logistic regression analyses, and the numbers and percentages (%) of positive cases in the PD and HC groups. The cut off value for age (*i*.*e*., 45.5 years of age) corresponds to the maximum Youden Index to best differentiate PD from HC subjects.n.a.=not available. PPMI=The Parkinson’s Progression Marker Initiative. PD=Parkinson disease. HC=healthy control. OR= odds ratio. CI=confidence interval. ROC=receiver operating characteristic. AUC=area under the ROC curve. RBD=REM Sleep Behavior Disorder. CSF=Cerebrospinal fluid.

### PPMI cohort

Several associations between selected variables and PD diagnosis, such as the presence of hyposmia, constipation, a positive family history of PD, genetic risk variants, depression, anxiety, RBD, as well as a reduction in CSF markers (for total α-synuclein and total tau) were statistically significant (**Figure 4**), as expected from the literature. In PPMI participants, hyposmia was particularly informative in distinguishing PD from HC (AUC: 0.82). Moreover, most variables showed similar diagnostic performances and directions of risk among the PPMI participants compared to the DeNoPa cohort, except for head trauma. Of note, in the PPMI cohort, several environmental exposure variables were not captured at the time of enrolment, which we had included as potentially informative in the original PREDIGT model.

Nevertheless, as illustrated in the score density plot (**Figure 5 (a)**), in the PPMI cohort, the HC group (n=173) had a lower PREDIGT Score compared with PD patients (n=374). The difference between mean scores (±SD) of these two groups was significant (p < 0.0001) at 47.19 (31.82) and 114.03 (54.69), respectively (**Figure 5(b)**). The AUC value for the PREDIGT Score using the PPMI cohort was 0.87 (95% CI 0.84-0.9) (**Figure 5 (c)**). Using 69.8 (indicated in **Figure 5 (a)** and **(b)** using red dashed lines) as the optimal threshold, the PREDIGT Score could distinguish PD patients from HC subjects in PPMI with 0.77 sensitivity (95% CI 0.73-0.81) and 0.82 specificity (95% CI 0.75-0.87).

**Figure 5:**
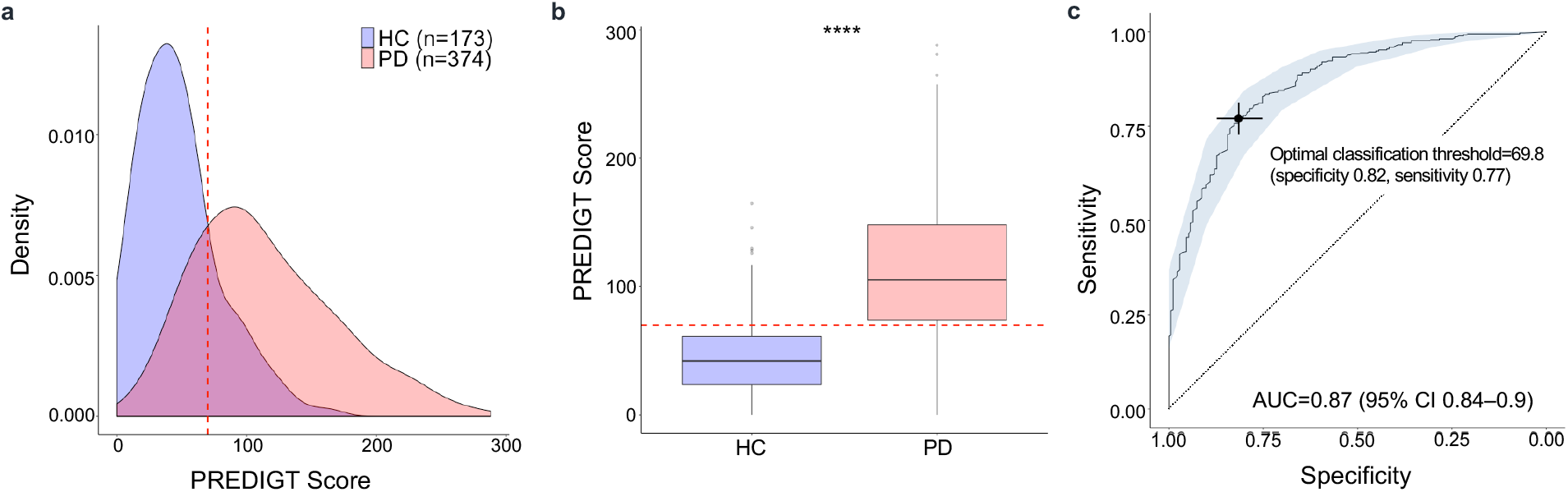
Cohort-specific evaluation of the PREDIGT Score performance using the PPMI cohort. (a) Score density plot; (b) box plot; and (c) ROC curve. The colour blue represents the HC group, pink represents PD patients. The mean score (with standard deviation) of healthy controls at 47.19 (±31.82) was significantly lower than that of the PD group at 114.03 (±54.69) (p<0.0001). Red, dashed lines in panels (a), (b) and the cross in (c) indicate the optimal threshold (score, 69.8) to distinguish between PD and HC groups corresponding to the maximum Youden Index (specificity, 0.82; sensitivity, 0.77). Light blue shading in the ROC curve indicates the bootstrap-estimated 95% CI within the AUC. DeNoPa=De Novo Parkinson. PD=Parkinson disease. HC=healthy controls. ROC=receiver operating characteristic. AUC=area under the ROC curve. CI=confidence interval.

### Shared variables model

With the utilization of fewer risk modifiers to compute a PREDIGT Score, we next tested the combined cohort, the DeNoPa study alone and the PPMI study alone. With only 11 variables included, the corresponding three AUC values were 0.84 (95% CI 0.81-0.87), 0.84 (95% CI 0.78-0.9) and 0.84 (95% CI 0.81-0.88), respectively (**Figure 6 (a)**). The mean PREDIGT Scores for PD patients and control individuals in the combined cohort with 11 shared variables were 108.48 (±52.08) and 47.33 (±34.1), respectively (p < 0.0001; **Figure 6 (b)**).

**Figure 6:**
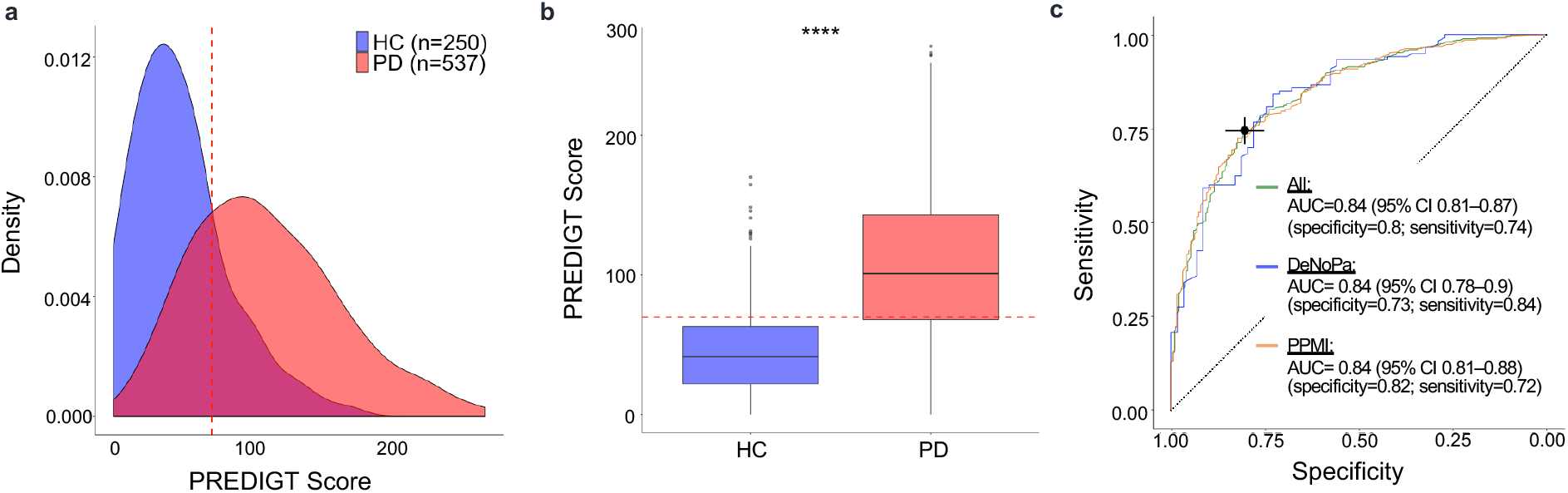
Evaluation of an eleven shared variables-based PREDIGT Score in the combined DeNoPa and PPMI cohorts. (a) Score density plot; (b) box plot; and (c) ROC curves. The colour blue represents the combined HC group, dark pink represents the combined PD group. The mean score (standard deviation) of healthy controls was significantly lower than that of the PD group at 47.33 (±34.1) and 108.48 (±52.08), respectively (p < 0.0001). Red, dashed lines in figure (a), (b) and cross in (c) represent the optimal threshold (69.8) that corresponds to the maximum Youden Index to differentiate HC from PD subjects in the combined cohort. Indicated sensitivity and specificity values in (c) are corresponding to AUC values for each individual study and the combined cohort based on PREDIGT Scores that uses eleven shared variables. DeNoPa=De Novo Parkinson. PPMI=The Parkinson’s Progression Marker Initiative. PD=Parkinson disease. HC=healthy controls. ROC=receiver operating characteristic. AUC=area under the ROC curve. CI=confidence interval.

Note, under the risk category ‘G’, males receive a higher value for sex (1.2) when computing a PREDIGT Score than females (0.8) (see **Suppl. Table 1**).^6^ Within each sex, the discriminative performance of the PREDIGT Score was generally similar for male and female subjects, as demonstrated by ROC curves and AUC values. As shown in **Suppl. Figure 1**, when we applied the formula to male subjects and female subjects separately, the results suggested that the PREDIGT Score was not sex biased, *i*.*e*., it didn’t perform significantly better for one sex than the other.

## DISCUSSION

Here, we assessed the performance of the original PREDIGT Score in two case-control cohorts. We found that this easy-to-use, formula-based scoring system discriminated PD patients from the HC group at the time of their enrolment with relatively high accuracy using readily accessible data and without the reliance on any results for motoric symptoms and signs (**Figures 3, 5, 6**).

The strengths and implications of this study are: (1) The performance of the PREDIGT Score was evaluated using two well characterized cohorts in which many (but not all) variables from the original score sheet had been captured. (2) The outcomes were similar for the single-center study (DeNoPa) *vs*. the multi-centric (PPMI) cohort. There, accuracy of the clinical diagnosis was achieved through annual follow up visits in both cohorts and DAT-scan imaging of all enrolled patients in PPMI, thereby permitting robust group classification with the elimination of false positives and reliable algorithm assessment. (3) The numerical values assigned to the chosen variables were based on published meta-analyses and had previously been reported by us; therefore, the model can be further tested using other cohorts and populations. (4) The fact that the *shared* model achieved similar AUCs with fewer variables, when compared to the *cohort-specific* evaluations of the PREDIGT Score, suggests that simplification of the original model could be explored for even greater ease of use in the future. (5) The current version of the PREDIGT Score sheet is largely based on information collected via validated questionnaires that need limited involvement by a licensed, clinical practitioner. Of note, the individual effect sizes of two CSF biomarkers, which require a relatively invasive procedure (*i*.*e*., lumbar puncture) and several analytical steps in the laboratory (*e*.*g*., ELISAs), were rather small. Thus, these two variables could theoretically be removed from a modified version (**Figures 2, 4**). It follows, that in the future the PREDIGT Score could be transformed into a screening tool and/or an online calculator to be completed in practitioners’ offices and/or by subjects at home. If further validated, the PREDIGT Score could thus be of diagnostic use in routine patient care as well as in clinical research studies during screening efforts and enrolment steps. (6) Last but not least, the ongoing evaluation of the PREDIGT Score’s performance in well-designed cohorts may inform the complex aetiology and pathogenetic evolution of typical PD. Conversely, newly emerging epidemiological and mechanistic insights into the development of PD will generate opportunities to better integrate additional risk elements into updated versions of the PREDIGT Score sheet, such as those associated with concrete infections and/or microbial colonization under category ‘E’, and those with evidence of dysregulated inflammation, as determined for example by cytokines measured in biological fluids, under category ‘I’.^16-20^

We are aware of the following limitations of this study and will address them in future work. Individuals enrolled in case-control studies are not always representative of the general population due to potential selection biases. Especially, the effect sizes of categories ‘G’ and ‘T’, two important risk modifiers in PD, were not adequately represented in DeNoPa and PPMI due to their sex-and age-matched study designs. Consequently, the performances of the PREDIGT Score reported here were rather conservative; therefore, the model’s power to distinguish between PD and HC subjects in other cohorts/populations, where sex and age are represented differently, would likely be higher. Because the predicted incidence of disease may be over-or under-estimated in any case-control cohort, we will also examine and calibrate coefficients of the PREDIGT Score in population settings during additional validation efforts.^21^ PREDIGT Scores reported here are raw scores and they only represent the relative risk (rank) between study participants within the DeNoPa and PPMI cohorts. Computation of scores that quantify the absolute risk of PD will require additional, external validation, especially population-level data.

The optimal cut-off values and related sensitivity/specificity results reported here correspond to the calculated maximum Youden Index. However, in clinical care settings, misclassified subjects (*i*.*e*., false positives and false negatives) may have different outcomes of monitoring, treatment and experimental interventions. Upon evaluation of the PREDIGT Score model in population settings, analyses that characterize and quantify implications for misclassified subjects will likely inform practice-related thresholds applied to clinical use.

DeNoPa and PPMI are two well established cohorts with their design having been informed by other studies.^22,23^ Still, there are variables for our analyses that were missing from both cohorts. For example, given the fact that CRFs in DeNoPa and PPMI had not yet included standardized questionnaires to capture subjects’ detailed medical history, select variables had to be eliminated from our study (**Figure 1**). Therefore, the results reported here represent an approximation.

With securing access to other cohorts’ data sets in the future, we plan to assess the PREDIGT Score’s discriminative performance in a similar, case-control manner (*e*.*g*., EPIPARK,^24^ Harvard Biomarker Study^22^). We will also undertake the task of assessing its predictive performance in prospective, longitudinal studies of neurologically healthy persons, who were enrolled after having been identified as carrying one or more risk factor, a key emphasis in our field, as highlighted by Noyce et al.^25^ There, we will seek to interrogate results from those ongoing, regional cohorts where sufficient data on variables needed for the calculation of the PREDIGT Score have been collected, such as in: PRIPS (Prospective validation of risk factors for the development of Parkinson syndromes);^26^ TREND (Tübingen-based evaluation of risk factors in the early detection of neurodegeneration);^27^ PARS (Parkinson At-Risk Syndrome);^28^ OPDC (Oxford Parkinson’s Disease Centre study);^29^ PREDICT-PD;^30^ Bruneck;^31^ and Luxembourg Parkinson’s Study.^32^

We also noticed that criteria that define concrete variables, which we impute into the PREDIGT Score sheet, may vary from cohort to cohort. For example, in their CRFs DeNoPa and PPMI employed different definitions of head trauma, which could explain its opposite effects on the association with the diagnosis of PD, although neither was statistically significant (**Figures 2, 4**). Different ELISA platforms had been used to quantify the total tau concentration in CSF, although the individual effect size of CSF tau concentration was also relatively small. Rigorously standardized CRFs and applied methodologies to quantify risk modification across all potential variables, in particular within the growing fields of exposome-, genome-, and biomarker-based research,^33^ will advance our abilities to update the PREDIGT Score and further interrogate it.

In accordance, variables currently captured by free text in the CRFs used in each study, such as those that relate to medical conditions and test results relevant to category ‘I’,^6^ need to be streamlined for subsequent quantification and inclusion. Capturing the detailed medical history of select chronic conditions, especially those linked to colonization by microbes and *bona fide* infections under category ‘E’ resulting in ongoing inflammation under ‘I’ (*e*.*g*., due to Crohn’s disease, hepatitis C and -B, as well as rosacea), are of great interest and will be included in later revisions of the model. Moreover, our current analysis focused on variables included in the original PREDIGT Score. Additional variables captured by CRFs in various cohorts, such as those related to other non-motor aspects, *e*.*g*., urinary dysfunction, sexual dysfunction, apathy, and fatigue, could also be tested and possibly included in future revisions. Furthermore, both the DeNoPa and PPMI cohorts employed multiple questionnaire-based data sheets some of which could be abbreviated to extract the most pertinent items, such as after applying Item Response Theory and other validated, psychometric methods (*e*.*g*., Optimal Scoring).^34^

Depending on the availability of appropriate cohorts, the model’s diagnostic and predictive performance model will also be tested on patients with other neurological diseases, including but not restricted to Alzheimer’s disease (AD), which shares hyposmia with PD as a common risk factor (or feature of a prodromal state),^35^ forms of dementia other than AD, as well as atypical parkinsonism and variants of secondary parkinsonism. We plan to evaluate the performance of the PREDIGT Score to distinguish PD patients *vs*. subjects that had ‘scans without evidence of a dopaminergic deficit’ (SWEDD); this, to better define its true discriminative power in a typical clinic setting, such as during the first 12-24 months after the onset of a limb tremor and change in muscle tone.

The currently assigned coefficients of PREDIGT variables are based on published meta-analysis and our initial hypothesis.^6^ Although our results demonstrate that the formula underlying the PREDIGT Score is relatively accurate in distinguishing PD from HC, the model’s performance could -in theory-also be improved by adjusting coefficients using statistical methods and available machine learning techniques. We have recently begun that work.

As envisioned, the ultimate goal of a fully validated PREDIGT Score is to identify at-risk individuals before the onset of parkinsonism and thus enable interventions aimed at reducing the incidence of PD. Three variables within the PREDIGT categories, namely ‘D’, ‘G’ and ‘T’, will not be amenable to interventions. However, cumulative exposure to protective agents, potentially harmful microbes, and neurotoxicants within the environment (‘E’) as well as select tissue responses to gene-environment interactions (‘I’), such as chronic inflammation and related dysregulation of α-synuclein outside and/or inside the central nervous system, could well represent targets for engagement in the future. Hence, a screening tool, such as a paper-based questionnaire and/or online calculator, will be developed and field tested to examine the utility of the PREDIGT Score in both clinic and home settings.

Last but not least, the successful validation of the PREDIGT Score, and possibly, our approach to deconstruct ‘idiopathic’ PD, could serve as a template for the aetiological modeling and mathematical interrogation of other complex syndromes in long-lived humans, such as coronary heart disease, diabetes and dementia.

## Data Availability

PPMI data used in the preparation of this article were obtained from the Parkinson's Progression Markers Initiative (PPMI) database (www.ppmi-info.org/data). For up-to-date information on the study, visit www.ppmi-info.org. For DeNoPa, please contact the study PI Dr. Brit Mollenhauer regarding data access.

## ACKNOWLEDGEMENTS

The authors acknowledge the commitment of study participants in the DeNoPa and PPMI cohorts and are grateful to all clinical research coordinators at their participating study sites. PPMI data used in the preparation of this article were obtained from the Parkinson’s Progression Markers Initiative (PPMI) database (www.ppmi-info.org/data). For up-to-date information on the study, visit www.ppmi-info.org. We thank Ms. Nathalie Lengacher for help in graphic design. We thank Dr. Anna Naito for help in data acquisition and coordination. This work was supported by funding from Parkinson Canada (to T.M., D. M., M.G.S; 2018; to J.L.; 2019-2021), Michael J. Fox Foundation for Parkinson’s Research (to T.M., D. M., M.G.S), Department of Medicine (T.M., T.R., D.M., M.G.S.), The Ottawa Hospital Foundation (Borealis Foundation to J.L.) and the Uttra & Subash Bhargava Family (M.G.S.), the Paracelsus-Elena-Klinik Kassel, Parkinson Fonds Deutschland, and the Deutsche Parkinson Vereinigung (B.M.; C.T.). PPMI - a public-private partnership - is funded by the Michael J. Fox Foundation for Parkinson’s Research and funding partners [list the full names of all of the PPMI funding partners found at www.ppmi-info.org/fundingpartners]. The funders had no role in the design and execution of the study; the collection, management, analysis, and interpretation of the data; the preparation, review, or approval of the manuscript; and the decision to submit the manuscript for publication. We are grateful for the ongoing support and feedback from people with lived experiences, such as through the board of the Parkinson Research Consortium Ottawa and members of Partners Investing in Parkinson’s Research, and to Drs. P. Wells and D. Lewis for their ongoing encouragement.

## AUTHORS’ CONTRIBUTIONS

JL, TAM, DM, and MGS contributed to the concept and design of the study. JL and MGS contributed to the acquisition of data. JL, TR, and DM decided on the statistical methods used in this study. JL did data cleaning, data analysis, figures and tables. JL, TAM, JJT, TR, DM, and MGS contributed to data interpretation. BM, MF, and CT contributed to the data collection and verification. JL, TAM, BM, MF, JJT, CT, TR, DM, and MGS contributed to the drafting of the article and revising it critically, and all approved the submission of current version.

---

**Supplementary Figure 1:**
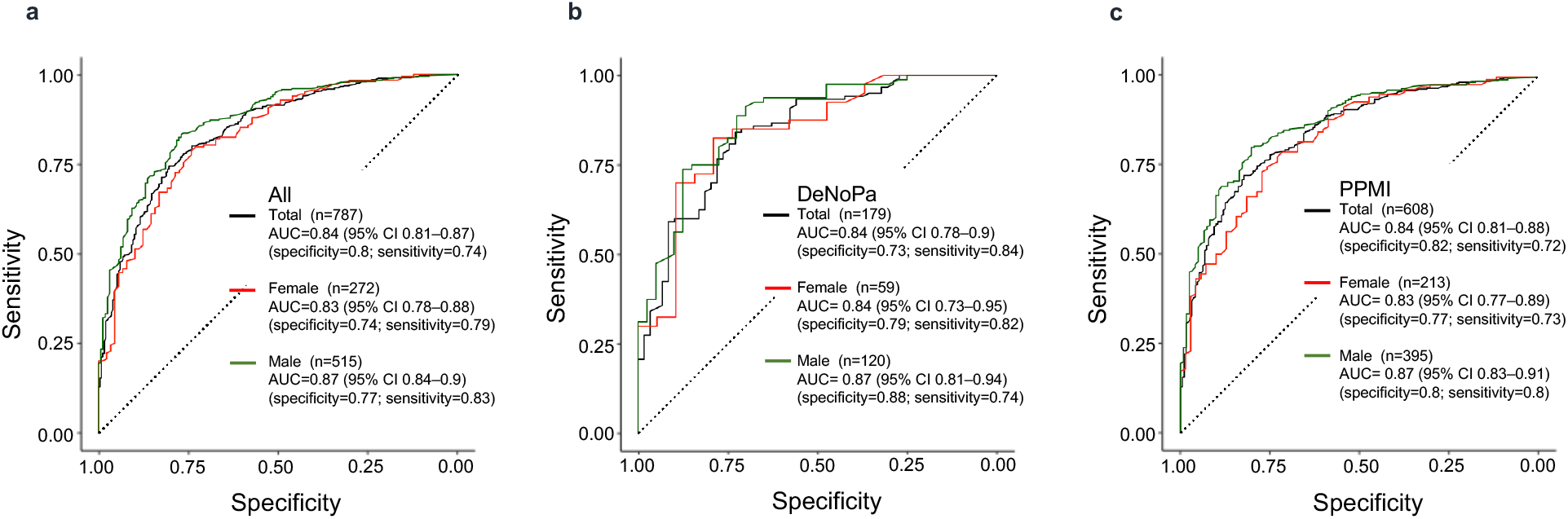
Probing for sex effects in the performance of an eleven shared variables-based performance of the PREDIGT Score in the DeNoPa and PPMI studies as well as the combined cohort. (a), (b), and (c) show ROC curves for the combined cohort, the DeNoPa study, and the PPMI cohort, respectively. Black curves represent all subjects, red curves represent female participants, and green curves represent male subjects. Indicated sensitivity and specificity values are corresponding to AUC values for each sex individually and both sexes taken together in the cohorts tested based on a PREDIGT Score model that uses eleven shared variables. DeNoPa=De Novo Parkinson. PPMI=The Parkinson’s Progression Marker Initiative. ROC=receiver operating characteristic. CI=confidence interval.

**Supplementary Table 1:**
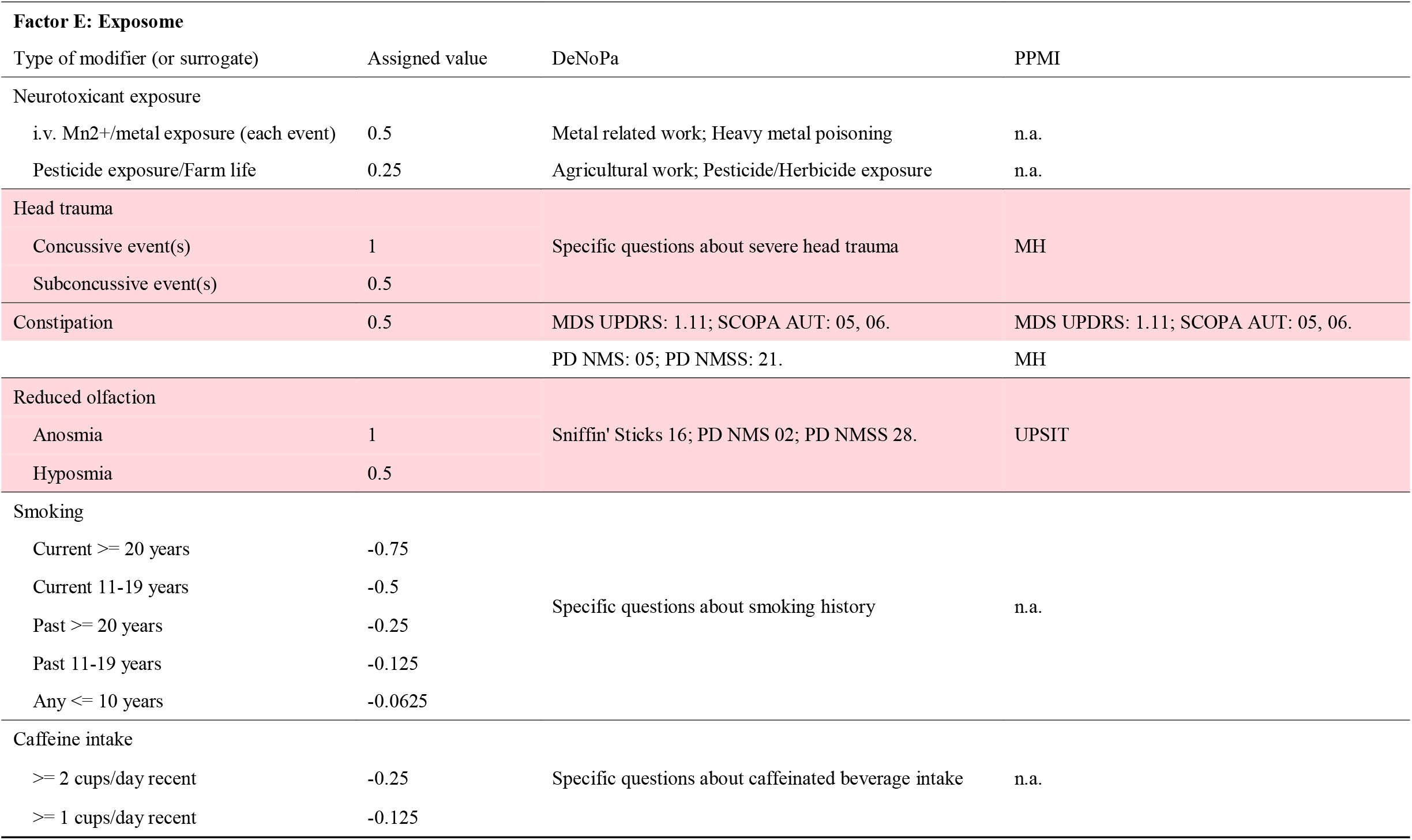
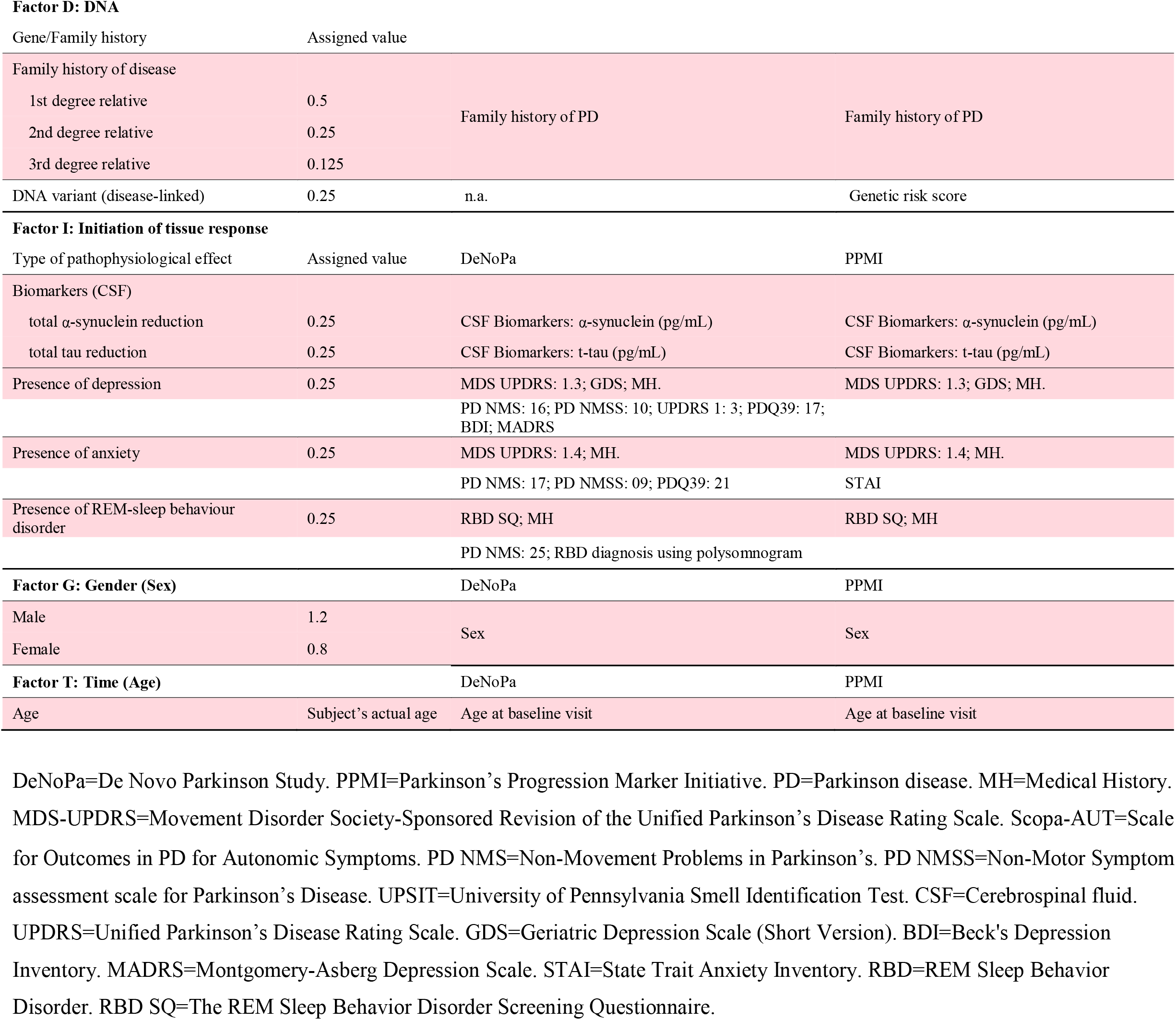
Select variables and their previously assigned values within five categories that are entered into the PREDIGT Score formula [PR=(E+D+I)xGxT)]. The eleven variables, which were captured in both DeNoPa and PPMI cohorts and included in the ‘shared evaluation’ of the PREDIGT Score (as shown in Figure 6) are shaded in pink.

**Supplementary Table 2:**
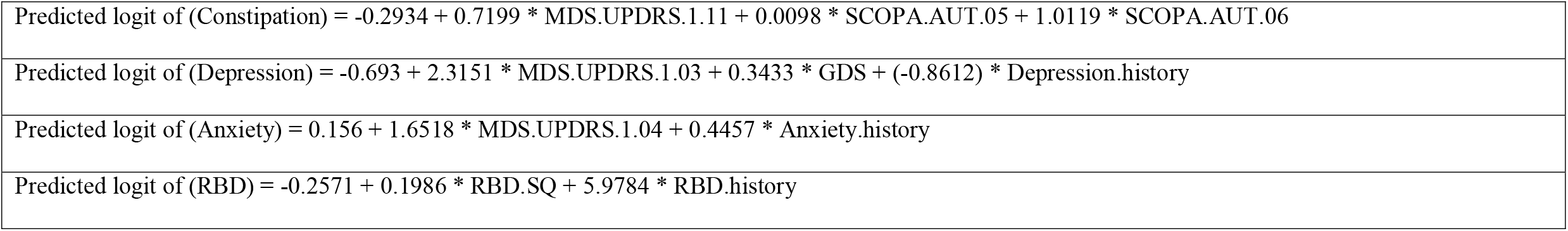
Summary scores for four variables generated by logistic regression analysis of multiple assessments-based data shared in the combined cohorts: constipation, depression, anxiety, and REM-sleep behaviour disorder (RBD).

## REFERENCES

1 Collaborators Global Burden of Disease (GBDPsD). Global, regional, and national burden of Parkinson’s disease, 1990□2016: a systematic analysis for the Global Burden of Disease Study 2016. Lancet Neurol 2018; 17: 939–953.

2 Dorsey ER, Constantinescu R, Thompson JP et al. Projected number of people with Parkinson disease in the most populous nations, 2005 through 2030. Neurology. 2007; 68: 384–386

3 Berg D, Godau J, Seppi K, et al. The PRIPS study: screening battery for subjects at risk for Parkinson’s disease. Eur J Neurol 2013; 20: 102–108

4 Nalls MA, McLean CY, Rick J, et al. Diagnosis of Parkinson’s disease on the basis of clinical and genetic classification: a population□based modelling study. Lancet Neurol. 2015; 14: 1002–1009.

5 Schrag A, Anastasiou Z, Ambler G, Noyce A, Walters K. Predicting diagnosis of Parkinson’s disease: A risk algorithm based on primary care presentations. Mov Disord 2019; 34(4): 480–486.

6 Schlossmacher MG, Tomlinson JJ, Santos G, et al. Modelling idiopathic Parkinson disease as a complex illness can inform incidence rate in healthy adults: the PREDIGT Score. Eur J Neurosci 2017; 45: 175–191.

7 Kalia LV, Lang AE. Parkinson’s disease. Lancet 2015; 386(9996): 896–912.

8 Mollenhauer B, Trautmann E, Sixel-Do□ring F, et al. Nonmotor and diagnostic findings in subjects with de novo Parkinson disease of the DeNoPa cohort. Neurology 2013; 81(14): 1226–34.

9 Parkinson Progression. The parkinson progression marker initiative (PPMI). Prog Neurobiol. 2011; 95: 629–635.

10 Hughes AJ, Daniel SE, Kilford L, Lees AJ. Accuracy of clinical diagnosis of idiopathic Parkinson’s disease. A clinico-pathological study of 100 cases. JNNP 1992; 55:181–184.

11 Rubin DB. Statistical Matching Using File Concatenation with Adjusted Weights and Multiple Imputations. Journal of Business & Economic Statistics 1986; 4 (1): 87–94.

12 Little R. Missing-Data Adjustments in Large Surveys. Journal of Business & Economic Statistics 1988; 6 (3): 287–296.

13 Hummel T, Kobal G, Gudziol H, Mackay-Sim A. Normative data for the “Sniffin’ Sticks” including tests of odor identification, odor discrimination, and olfactory thresholds: an upgrade based on a group of more than 3,000 subjects. Eur. Arch. Otorhinolaryngol 2007; 264: 237–243.

14 Muirhead N, Benjamin E, Saleh HA. Is the University of Pennsylvania Smell Identification Test (UPSIT) valid for the UK population? Otorhinolaryngologist 2012; 6(2): 99–103

15 Youden WJ. Index for rating diagnostic tests. Cancer. 1950 Jan;3(1):32–5.

16 Egeberg A, Hansen PR, Gislason GH, Thyssen JP. Exploring the Association Between Rosacea and Parkinson Disease: A Danish Nationwide Cohort Study. JAMA Neurol. 2016;73(5):529–534.

17 Hui KY, Fernandez-Hernandez H, Hu J, Schaffner A, Pankratz N, Hsu NY, et al.Functional variants in the LRRK2 gene confer shared effects on risk for Crohn’s disease and Parkinson’s disease. Sci Transl Med, 2018;10(423):eaai7795.

18 Tsai H-H, Liou H-H, Muo C-H, Lee C-Z, Yen R-F, Kao C-H. Hepatitis C virus infection as a risk factor for Parkinson disease: A nationwide cohort study. Neurology 2016, 86 (9):840–846.

19 Eidson LN, Kannarkat GT, Barnum CJ et al. Candidate inflammatory biomarkers display unique relationships with alpha-synuclein and correlate with measures of disease severity in subjects with Parkinson’s disease. J Neuroinflammation 2017;14;164.

20 Shutinoski B, Hakimi M, Harmsen IE, Lunn M, Rocha J, Lengacher N, et al. Lrrk2 alleles modulate inflammation during microbial infection of mice in a sex-dependent manner. Science Translational Medicine 2019; 11(511)

21 Manuel DG, Perez R, Sanmartin C, Taljaard M, Hennessy D, Wilson K, Tanuseputro P, Manson H, Bennett C, Tuna M, Fisher S, Rosella LC. Measuring Burden of Unhealthy Behaviours Using a Multivariable Predictive Approach: Life Expectancy Lost in Canada Attributable to Smoking, Alcohol, Physical Inactivity, and Diet. PLoS Med. 2016; 13(8): e1002082.

22 Ding H, Dhima K, Lockhart KC, Locascio JJ et al. Unrecognized vitamin D3 deficiency is common in Parkinson disease. Harvard Biomarker Study. Neurology Oct 2013; 81 (17): 1531–1537

23 Locascio JJ, Eberly S, Liao Z, Liu G, Hoesing AN, Duong K, Trisini-Lipsanopoulos A, Dhima K, Hung AY, Flaherty AW, Schwarzschild MA, Hayes MT, Wills AM, Shivraj Sohur U, Mejia NI, Selkoe DJ, Oakes D, Shoulson I, Dong X, Marek K, Zheng B, Ivinson A, Hyman BT, Growdon JH, Sudarsky LR, Schlossmacher MG, Ravina B, Scherzer CR. Association between α-synuclein blood transcripts and early, neuroimaging-supported Parkinson’s disease. Brain 2015; 138(Pt 9): 2659–2671.

24 Kasten M, Hagenah J, Graf J, et al. Cohort Profile: a population-based cohort to study non-motor symptoms in parkinsonism (EPIPARK). Int J Epidemiol 2013; 42: 128–128k.

25 Noyce AJ, Lees AJ, Schrag A, The prediagnostic phase of Parkinson’s disease. Journal of Neurology, Neurosurgery & Psychiatry 2016; 87: 871–878.

26 Berg D, Seppi K, Behnke S, et al. Enlarged substantia nigra hyperechogenicity and risk for Parkinson disease: a 37-month 3-center study of 1847 older persons. Arch Neurol 2011; 68: 932–7.

27 Gaenslen A, Wurster I, Brockmann K, et al. Prodromal features for Parkinson’s disease— baseline data from the TREND study. Eur J Neurol 2014; 21: 766–72

28 Jennings D, Siderowf A, Stern M, et al. Imaging prodromal Parkinson disease: the Parkinson Associated Risk Syndrome study. Neurology 2014; 83:1739–1746.

29 Baig F, Lawton M, Rolinski M, et al. Delineating nonmotor symptoms in early Parkinson’s disease and first-degree relatives. Mov Disord. 2015; 30: 1759–1766.

30 Noyce AJ, Bestwick JP, Silveira-Moriyama L, et al. PREDICT-PD: Identifying risk of Parkinson’s disease in the community: methods and baseline results. Journal of Neurology, Neurosurgery & Psychiatry 2014; 85: 31–37.

31 Marini K, Mahlknecht P, Tutzer F, Stockner H, Gasperi A, Djamshidian A, et al. (2020), Application of a Simple Parkinson’s Disease Risk Score in a Longitudinal Population□Based Cohort. Mov Disord 2020; 35: 1658–1662.

32 Hipp G., Vaillant M., Diederich N.J., Roomp K., Satagopam V.P., Banda P., et al.The Luxembourg Parkinson’s Study: A Comprehensive Approach for Stratification and Early Diagnosis. Frontiers in Aging Neuroscience 2018; 10:326.

33 Mollenhauer, B. et al. Biological confounders for the values of cerebrospinal fluid proteins in Parkinson’s disease and related disorders. J. Neurochem. 2016; 139 (Suppl. 1), 290–317.

34 Ramsay JO, Wiberg M, Li J. Full Information Optimal Scoring. Journal of Educational and Behavioral Statistics 2019; 45(3): 297–315

35 Sun GH, Raji CA, Maceachern MP, Burke JF. Olfactory identification testing as a predictor of the development of Alzheimer’s dementia: a systematic review. Laryngoscope. 2012 Jul; 122(7): 1455–62.

